# Labour market participation after a sickness absence due to cancer: a dynamic cohort study in Catalonia (Spain)

**DOI:** 10.1101/2022.12.01.22282934

**Authors:** Amaya Ayala-Garcia, Fernando G. Benavides, Laura Serra

## Abstract

**Background:** Consequences of cancer on working life until retirement age remain unclear. This study aimed to compare labour market participation patterns in workers with a sickness absence (SA) due to cancer versus those with no SA and those with SA due to other diseases.

**Methods:** Registry-based cohort study of social security affiliates in Catalonia from 2012-2018. Cases consisted of workers with SA due to cancer between 2012-2015 (N=516) and were individually age- and sex-matched with an affiliate with SA due to other diagnoses and a worker without SA. All workers (N=1,548, 56% women) were followed-up until the end of 2018. Sequence analysis, optimal matching, and multinomial logistic regression were used to identify and assess the probability of future labour market participation patterns (LMPP). All analyses were stratified by sex.

**Results:** Compared with workers with SA due to cancer, male workers without SA and SA due to other causes showed lower probability of being in the LMPP of death (aRRR 0.02, 95% CI: 0.00−0.16; aRRR 0.17, 95% CI: 0.06−0.46, respectively), and in women lower probability of increasing permanent disability and death (aRRR 0.24, 95% CI: 0.10−0.57; aRRR 0.39, 95% CI: 0.19−0.83). Compared to workers with SA due to cancer, risk of future retirement was lower in workers with no SA (women aRRR 0.60, 95% CI: 0.22−1.65; men aRRR 0.64, 95%CI: 0.27−1.52).

**Conclusions:** Workplaces should be modified to the needs of cancer survivors in order to prevent more frequent early exit of labour market due to retirement and permanent disability when possible.

**Key Messages:**

- **What is already known on this topic**. After the treatment stage and sickness absence (SA) period, some cancer survivors face adverse effects that affect their long-term work capacity and increase the likelihood of an early exit from labour market.
- **What this study adds** Labour market participation after cancer show that workers with an SA due to cancer manage to return to work and have stable employment. Anyhow, they show a higher likelihood of early retirement, receiving permanent disability benefits, and of dying than workers without a previous SA.
- **How this study might affect research, practice or policy** Cancer survivors require their new health status to be considered when return to work. Actions should be taken in order to regulate programmes that help them remain working when possible and desired.

## INTRODUCTION

In 2020, 385 new cases of cancer per 100,000 persons aged 20-64 years were diagnosed in Europe [1]. This figure represents nearly 50% of the total number of new cancer diagnoses [2], and the average 5-year survival of malignant neoplasms considering all ages has reached almost 54% [3]. A recent report predicted an extension of working life, with a 10% increase in the participation rate among workers aged between 64 and 74 years by 2070 in the EU-27 [4]. Therefore, an increase in the number of people diagnosed with cancer while working is expected.

Currently, most cancer survivors take a sickness absence (SA) during their treatment to overcome the acute stage of the disease [5] with the intention of returning to work when treatment ends. These SAs tend to be longer than SA spells due to other diagnoses [6]. After the treatment stage, some cancer survivors face adverse effects that can persist for prolonged periods or become chronic due to the treatment or the severity of the disease itself [7]. When these symptoms impair work performance, cancer survivors may decide to ask for permanent disability (PD) benefits from the social security system. The process of returning to work or not is also affected by other factors such as age, sex, family support, work and employment conditions, job strain, physical job demands, type of job, support at the workplace, type of contract and previous periods of unemployment [8–10]. However, most cancer survivors attempt to return to work (RTW) after the first year or a maximum of 2 years after diagnosis, right after the SA ends [11].

Literature on long-term working life, including all the possible working paths after cancer, is scarce. Most studies look at the probabilities of future labour outcomes individually, which obviates transitions between these states until retirement. Characterising the working paths of cancer survivors may shed light on how surviving the disease and subsequent career decisions may interact in the long term. In a previous study, we showed that cancer survivors are less likely to accumulate days of employment in the long term [12]. On the one hand, these changes in survivors’ working life may be driven by personal decisions due to health or financial status or a desire to modify career paths after a reassessment of life priorities [13]. Hence, cancer survivors may decide to work fewer hours than before the SA, take time off from work for prolonged periods, and experience cancer-associated long- or short-term job loss with or without unemployment benefits [14]. In addition, some survivors choose to change their retirement plan by retiring early or before planned [14]. On the other hand, these decisions could also be driven by labour market or workplace demands. For example, some survivors may not be able to carry out a high-strain job due to long-term side effects impairing their ability to work [15], and there may be a lack of appropriate jobs for their new health status. These decisions may take place at different points after RTW and combine differently depending on opportunities in the labour market and workplace factors. The negative impact of cancer on working life could increase inequalities that could and should be addressed to lessen financial and psychological consequences.

We hypothesised that cancer survivors would have more unstable career paths after returning to working activity and have a higher probability of prematurely leaving the labour market than people with other diseases or the general working population (Figure 1).

The objective of this study was to analyse all possible labour market states in a sample of workers after an SA due to cancer and to compare their working life paths to those of a sample of workers without SA and to workers with an SA due to other diseases.

**Figure 1.**
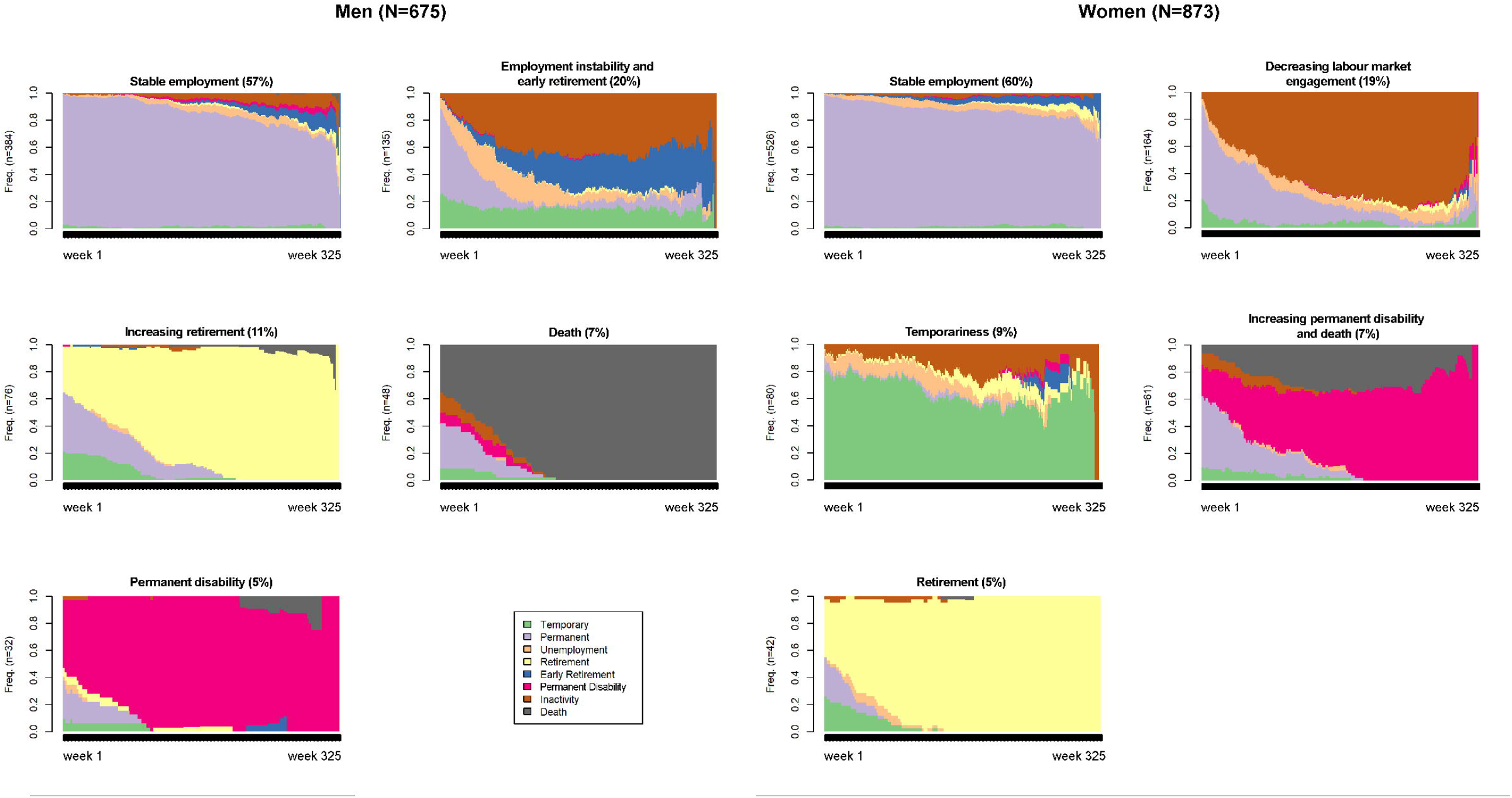
SEQ Figure \* ARABIC 1 Directed acyclic graph (DAG) of the relationship between having a sickness absence due to cancer and future working life. *categories of the variable “Return to work t_1_”: temporary employment, umemployment, permanent employment; * *categories of the variable “Return to work t_2_”:permanent disability, early retirement, retirement, inactivity and premature death.

## METHODS

Data were extracted from the Continuous Working Life Sample, which, since 2004, has taken an annual random representative sample of 4% of affiliates of the Spanish social security system. This database contains a full employment history of each affiliate, including information such as occupational category, the company’s economic activity, employment conditions (e.g., type of contract, income, and working time), social benefits (e.g., unemployment, PD, and retirement), other work-related variables (e.g., company ownership and size), and date of death. Based on this database, we were able reconstruct working life in the Spanish WORKss cohort [16].

In addition, in this study, we linked the database of the Catalan Institute for Medical and Health Evaluations, from which we obtained information on SA records between 2012 to 2015, including the medical diagnosis of the episode coded according to the 10th edition of the International Classification of Diseases (ICD-10), as well as the start and end date [17] of workers affiliated to the Spanish social security system in Catalonia, who were also part of the Spanish WORKss cohort.

We performed a registry-based cohort study among 1548 salaried workers living in Catalonia (675 men and 873 women). The study sample included salaried workers who had had an SA due to a malignant neoplasm (ICD-10, C00-C97) between 2012 and 2015 (N=516, 225 men and 291 women), and two age- (within a 5-year range) and sex-matched comparison groups with salaried workers with an SA due to a medical diagnosis other than cancer (n=516), and salaried worker without an SA (n=516). These participants were selected the week that the SA due to cancer ended (i.e., similar onset of time at risk) (Supplementary Table 1 and supplementary Table 2).

The average age of the three comparison groups in 2012 was 49.8 years for men (standard deviation: 9.96) and 47.0 years for women (standard deviation: 9.44). Among the comparison group of workers with an SA diagnosis other than cancer, the most frequent cause was diseases of the musculoskeletal system and connective tissue in both men and women (Supplementary table 2)

Each worker’s working life was characterised weekly from the time they entered the cohort between 2012 and 2015 until December 31, 2018, according to the possible labour states after an SA due to cancer. The study period ranged from 3 to 7 years, and the time out of the cohort was censored. The possible states were: temporary employment, permanent employment, unemployment with benefits, inactivity (considered as not having contact with social security longer than 15 days), PD (including total, absolute and severe degrees), early retirement, ordinary retirement, and death. If a worker was in two different states in the same week, that worker was assigned the state where he/she spent the longer time.

Potential confounders included in our analysis were occupational category (non-manual skilled, non-manual non-skilled, manual skilled, or manual non-skilled); working time categorised as a percentage of weekly hours (full-time [>87.5%], part-time [50%-87.5%], or short and marginal part-time [≤37.5%-49%]); monthly average income in tertiles (high [>€2370], medium [€1450 – 2370], or low [≤€1450]); company size (small/medium [up to 100 employees] and big [>100 employees]); company ownership (private and public); and the company’s economic activity (primary sector [agriculture, hunting, forestry, fishing, mining, and quarrying]; manufacturing [including construction and energy], and services). We also considered the previous 5-year employment ratio expressed as a percentage of employed days to the total potential working days, including working or unemployed or not affiliated days, to assess attachment to the labour market before the SA due to cancer. Workers who changed categories over time were assigned the category in which they spent most of the follow-up period.

Patients were not involved in any stage of the study, and confidentiality was maintained in both databases. The authors received data that were previously anonymised.

### Statistical analysis

The sample was described according to the above-mentioned response variables, explanatory variables and covariates, and the chi-square test was applied to assess the significance between comparison groups (Supplementary table 1). Sequence analysis was performed to reconstruct individual working lives by generating an ordered list of weekly labour states after an SA due to cancer. Optimal matching and cluster analyses were applied to identify groups of workers sharing similar working life trajectories. We called the resulting trajectories future labour market participation patterns (LMPP). Average silhouette width (ASW) was used to select the optimal number of clusters (values higher than 0.51 are recommended; Supplementary table 3) [18,19].

To measure the association between having an SA due to cancer and future LMPPs versus the comparison groups, we applied multinomial logistic regression with its relative risk ratio (RRR) and 95% confidence interval (95%CI).

All analyses were stratified by sex. Stata v.13 software was used for multinomial regression models, and R v.4.1.0 was used for sequence analysis and optimal matching analysis.

### Ethics approval and consent to participate

This study was performed in accordance with the standards of Good Clinical Practice and the principles of the Declaration of Helsinki. The study protocol guaranteed the fulfilment of Regulation (EU) 2016/679 of the European Parliament and the Council of 27 April 2016 on the protection of natural persons regarding the processing of personal data and the free movement of such data. It also fulfilled Spanish Organic Law 3/2018 of 5 December on the Protection of Personal Data and the Guarantee of Digital Rights. This study was approved by the Parc de Salut Mar Ethics Committee in Barcelona (Research Protocol no. 2020/9119) and was exempted from informed consent requirements owing to its registry-based design. The research team committed itself to the strict use of data for the present study. In addition, a linkage protocol agreement between the Centre for Research in Occupational Health at Pompeu Fabra University, the National Social Security Institute, and the Catalonian Institute for Medical Evaluations guaranteed the maintenance of confidentiality in providing the identified datasets to the authors.

## RESULTS

We found five LMPPs in both sexes. Among women, shared LMPPs were stable employment (60.3%), decreasing labour market engagement (18.7%), temporariness (9.1%), increasing PD and death (7.0%), and retirement (4.8%). Men’s future LMPP were summarised in stable employment (56.9%), employment instability and early retirement (20.0%), increasing retirement (11.4%), death (7.1%), and PD (4.7%) (Figure 2, Table 1).

**Figure 2.**
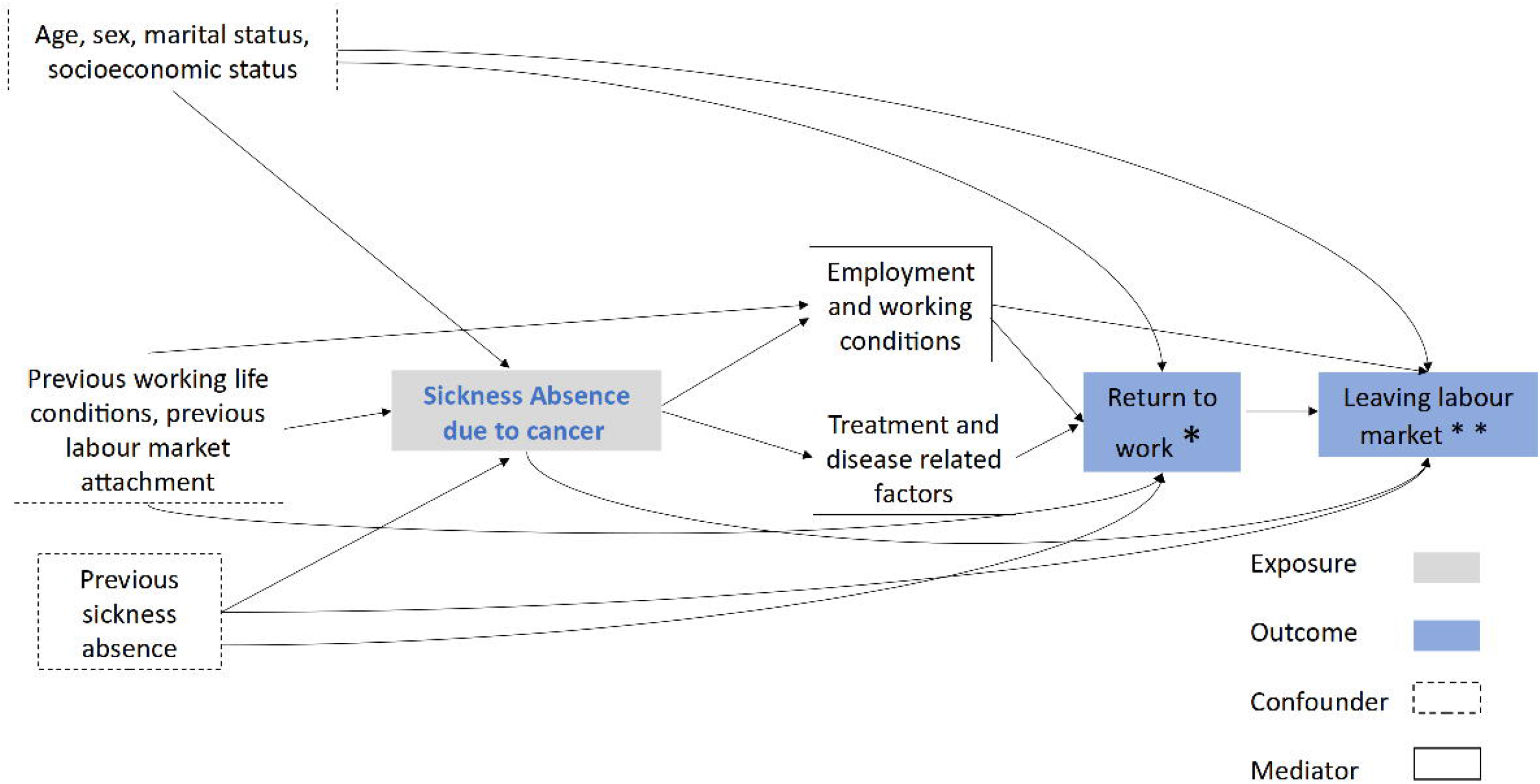
Future labour market participation patterns (LMPPs) in a sample of salaried men (top) and women (bottom) in Catalonia (2012–2018)

**Table 1.**
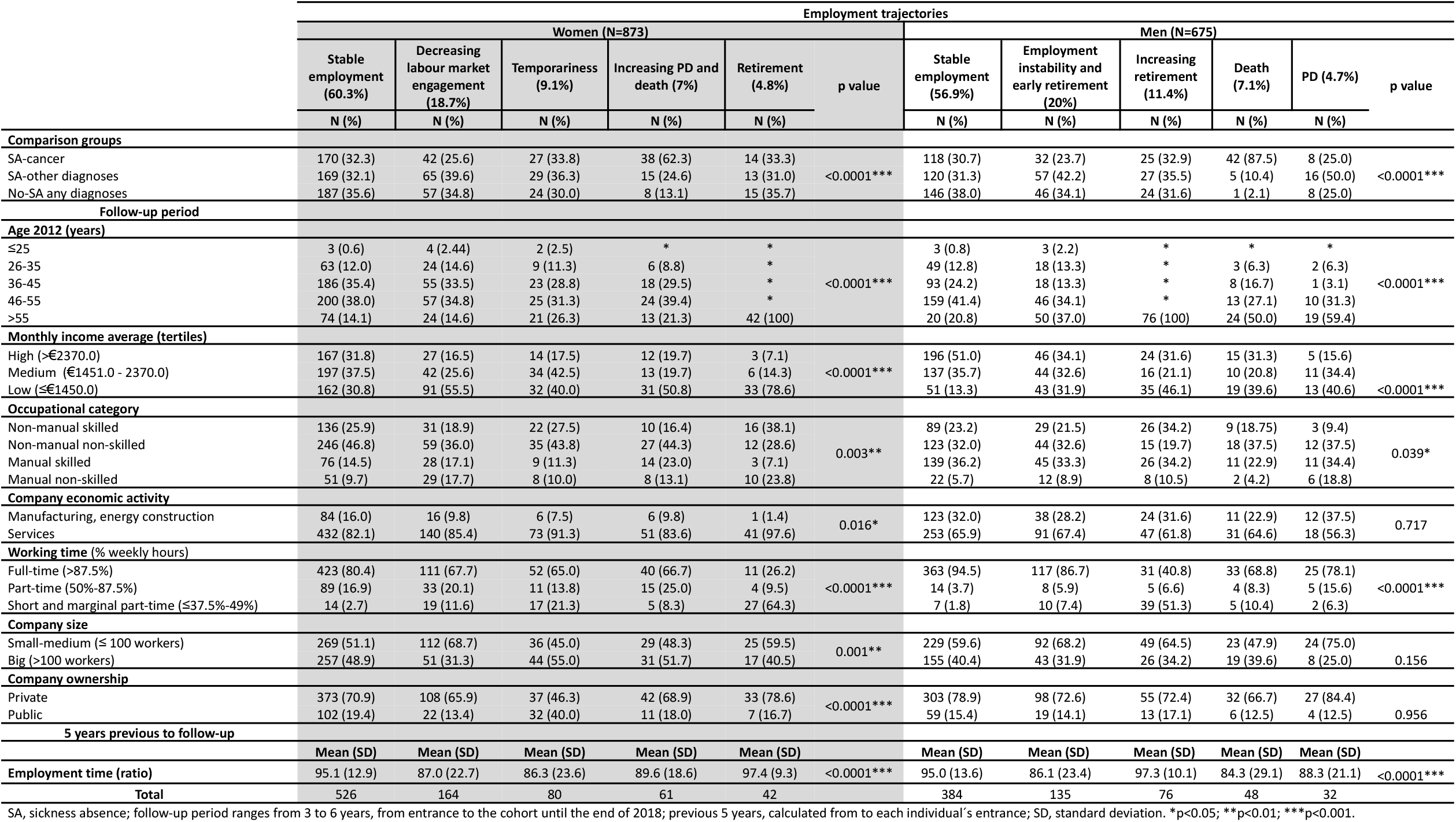
Future labour market participation patterns (LMPPs) in salaried workers living in Catalonia (2012–2018).

In both sexes, the most frequent LMPP was stable employment. Among workers with an SA due to cancer, 32.3% of women and 30.7% of men were in this pattern (Table 1). In this LMPP, 80% of workers were employed on a permanent contract throughout the follow-up period (Figure 2), and monthly income tended to be high (31.8% of women and 51.0% of men). In women, it also showed the lowest proportion of manual non-skilled occupations (9.7%) (Table 1).

Taking this stable employment LMPP as the reference category in both sexes, and compared with workers with an SA due to cancer, we examined the probability of belonging to each LMPP among workers with an SA due to other causes and workers without an SA (Tables 2 and 3). Compared with the SA due to cancer reference group, workers with an SA due to other causes had a higher probability of being in a low engagement, employment instability and early retirement LMPP rather than in a stable employment LMPP (men aRRR 1.76, 95% CI: 1.00−3.11; women aRRR 1.72, 95% CI: 1.02−2.90) (Table 2). This LMPP represented the second most frequent LMPP in men and women, showing a group of workers mostly employed on permanent contracts (60.0%), but very soon they started switching to unemployment benefits and inactivity, or to early retirement, particularly men (Figure 2).

**Table 2.** Association between future labour market participation patterns (LMPPs), company characteristics, and employment conditions among female salaried workers with a SA due to cancer (reference) and comparison groups.

|  | Women |  |  |  |  |  |  |  |  |  |  |  |
| --- | --- | --- | --- | --- | --- | --- | --- | --- | --- | --- | --- | --- |
|  | Decreasing labour market engagement vs stable employment |  |  | Temporariness vs stable employment |  |  | Retirement vs stable employment |  |  | Increasing PD and death vs stable employment |  |  |
|  | RRR | 95% CI | p value | RRR | 95% CI | p value | RRR | 95% CI | p value | RRR | 95% CI | p value |
| **Crude** |  |  |  |  |  |  |  |  |  |  |  |  |
| SA-cancer | 1 |  |  | 1 |  |  | 1 |  |  | 1 |  |  |
| SA-other diagnoses | 1.56 | (1.00–2.42) | 0.360 | 1.08 | (0.61–1.90) | 0.789 | 0.93 | (0.43–2.08) | 0.865 | 0.40 | (0.21–0.75) | 0.004** |
| No-SA any diagnoses | 1.23 | (0.78–1.93) | 0.050 | 0.81 | (0.45–1.45) | 0.477 | 0.97 | (0.29–2.22) | 0.946 | 0.19 | (0.09–0.42) | <0.0001*** |
| **Individually adjusted by:** |  |  |  |  |  |  |  |  |  |  |  |  |
| **Income (tertiles)** |  |  |  |  |  |  |  |  |  |  |  |  |
| SA-other diagnoses | 1.55 | (0.98–2.45) | 0.059 | 1.06 | (0.60–1.87) | 0.850 | 0.92 | (0.41–2.05) | 0.831 | 0.42 | (0.22–0.83) | 0.012* |
| No-SA any diagnoses | 1.12 | (0.70–1.78) | 0.630 | 0.79 | (0.43–1.42) | 0.425 | 0.84 | (0.38–1.82) | 0.650 | 0.20 | (0.09–0.45) | <0.0001*** |
| **Occupational category** |  |  |  |  |  |  |  |  |  |  |  |  |
| SA-other diagnoses | 1.47 | (0.92–2.36) | 0.111 | 1.08 | (0.60–1.94) | 0.808 | 1.01 | (0.45–2.25) | 0.983 | 0.32 | (0.16–0.62) | 0.001** |
| No-SA any diagnoses | 1.21 | (0.75–1.95) | 0.435 | 0.77 | (0.42–1.43) | 0.414 | 0.96 | (0.44–2.11) | 0.926 | 0.18 | (0.08–0.40) | <0.0001*** |
| **Company economic activity** |  |  |  |  |  |  |  |  |  |  |  |  |
| SA-other diagnoses | 1.67 | (1.06–2.64) | 0.027* | 1.11 | (0.62–1.96) | 0.725 | 1.03 | (0.47–2.26) | 0.944 | 0.41 | (0.21–0.80) | 0.009** |
| No-SA any diagnoses | 1.37 | (0.86–2.17) | 0.185 | 0.87 | (0.48–1.57) | 0.643 | 1.07 | (0.50–2.29) | 0.862 | 0.22 | (0.10–0.49) | <0.0001*** |
| **Working time (% weekly hours)** |  |  |  |  |  |  |  |  |  |  |  |  |
| SA-other diagnoses | 1.58 | (1.01–2.47) | 0.047* | 1.03 | (0.58–1.83) | 0.923 | 0.80 | (0.33–1.92) | 0.610 | 0.41 | (0.23–0.77) | 0.006** |
| No-SA any diagnoses | 1.26 | (0.80–1.99) | 0.321 | 0.78 | (0.43–1.42) | 0.410 | 0.86 | (0.37–2.03) | 0.732 | 0.20 | (0.09–0.44) | <0.0001*** |
| **Company size** |  |  |  |  |  |  |  |  |  |  |  |  |
| SA-other diagnoses | 1.68 | (1.07–2.64) | 0.023* | 1.06 | (0.60–1.87) | 0.834 | 0.96 | (0.44–2.10) | 0.914 | 0.40 | (0.21–0.77) | 0.005* |
| No-SA any diagnoses | 1.24 | (0.79–1.96) | 0.352 | 0.81 | (0.45–1.46) | 0.490 | 0.97 | (0.45–2.06) | 0.929 | 0.20 | (0.09–0.44) | <0.0001*** |
| **Company ownership** |  |  |  |  |  |  |  |  |  |  |  |  |
| SA-other diagnoses | 1.65 | (1.01–2.71) | 0.045* | 1.14 | (0.62–2.12) | 0.675 | 1.00 | (0.45–2.23) | 0.999 | 0.41 | (0.21–0.81) | 0.010* |
| No-SA any diagnoses | 1.35 | (0.82–2.24) | 0.242 | 0.93 | (0.49–1.77) | 0.826 | 1.04 | (0.47–2.27) | 0.931 | 0.24 | (0.11–0.54) | 0.001** |
| **Previous 5-year employment time (ratio)** |  |  |  |  |  |  |  |  |  |  |  |  |
| SA-other diagnoses | 1.62 | (1.03–2.54) | 0.036* | 1.13 | (0.64–2.00) | 0.683 | 0.92 | (0.42–2.02) | 0.833 | 0.41 | (0.22–0.77) | 0.006* |
| No-SA any diagnoses | 1.26 | (0.80–2.00) | 0.320 | 0.81 | (0.44–1.47) | 0.488 | 0.97 | (0.45–2.07) | 0.937 | 0.20 | (0.09–0.43) | <0.0001*** |
| **Adjustment by all variables:** |  |  |  |  |  |  |  |  |  |  |  |  |
| SA-other diagnoses | 1.72 | (1.02–2.90) | 0.043* | 1.09 | (0.56–2.13) | 0.801 | 0.72 | (0.26–1.95) | 0.515 | 0.39 | (0.19–0.83) | 0.014* |
| No-SA any diagnoses | 1.23 | (0.72–2.11) | 0.442 | 0.90 | (0.45–1.80) | 0.762 | 0.60 | (0.22–1.65) | 0.324 | 0.24 | (0.10–0.57) | 0.001** |

SA, sickness absence; PD, permanent disability; RRR, relative risk ratio; 95% CI, 95% confidence intervals. \*p<0.05; \*\*p<0.01; \*\*\*p<0.001.

**Table 3.**
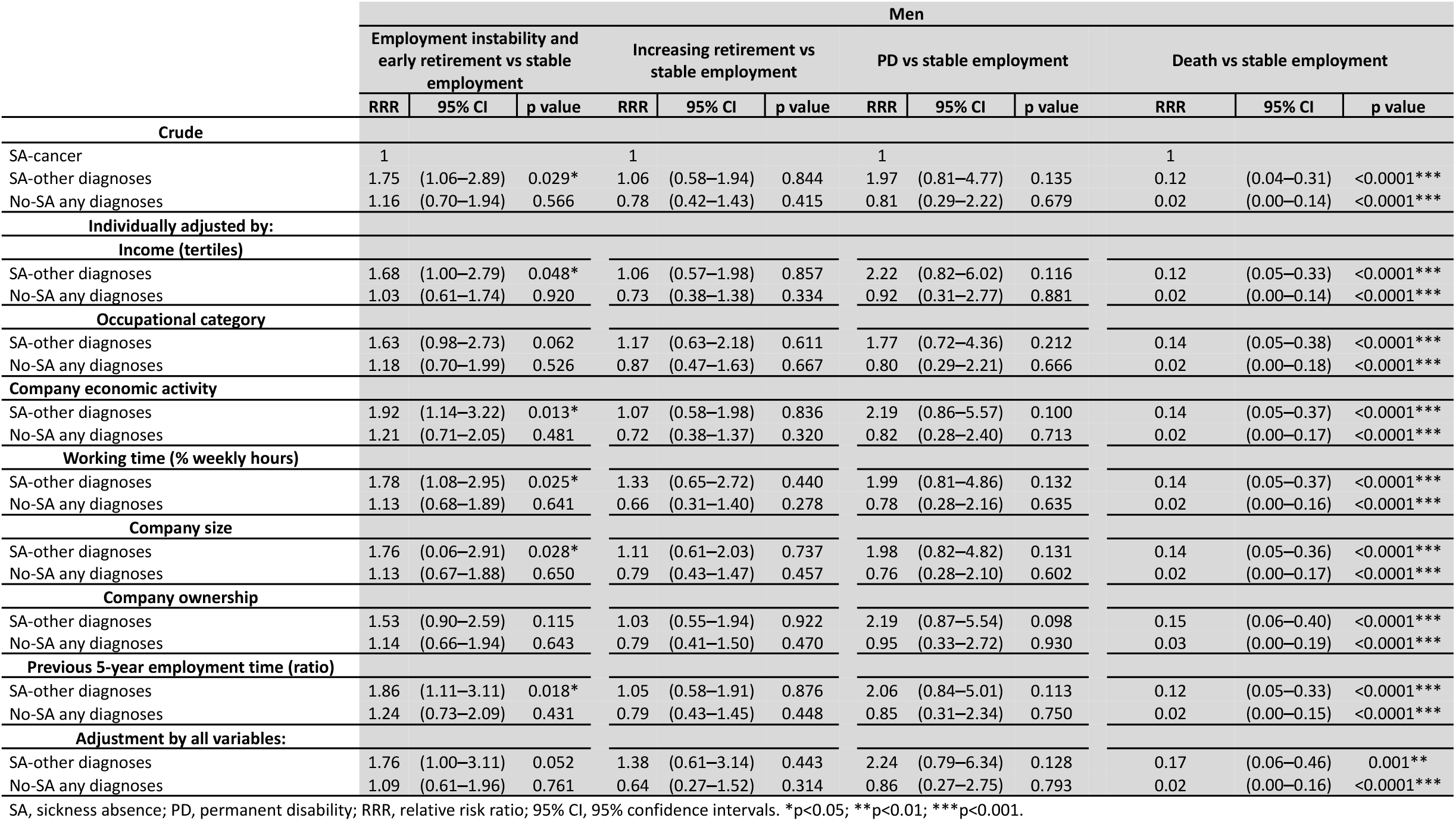
Association between future labour market participation patterns (LMPPs), company characteristics, and employment conditions among male salaried workers with a SA due to cancer (reference) and comparison groups.

Compared with workers with an SA due to cancer, both comparison groups showed a lower probability of death in men (no SA aRRR 0.02, 95% CI: 0.00−0.16; SA due to other causes aRRR 0.17, 95% CI: 0.06−0.46), and increasing PD and death LMPP in women (no SA aRRR 0.24, 95% CI: 0.10−0.57; SA due to other causes aRRR 0.39, 95% CI: 0.19−0.83). In men, these patterns differed from those in women (Table 2 and 3).

The pattern showing workers on PD was more frequent among men with an SA due to other causes (50.0%), and those aged over 55 years (55.6%). The pattern depicting workers who died during follow-up consisted mainly of men with an SA due to cancer (Table 1).

In women, unlike in men, an LMPP of temporariness represented 9% of the sample (Figure 2). Women without an SA showed trends towards a lower likelihood of being in a temporariness pattern (aRRR 0.90, 95% CI: 0.45−1.80) (Table 3). This LMPP showed the lowest percentage of employment in the 5 years before entering the cohort (86.3%) (Table 1).

When we assessed the probability of having an LMPP of exit from the labour market due to retirement, workers without an SA showed trends of a lower likelihood of being in this pattern, especially when adjusted by all employment and working conditions (women aRRR 0.60, 95% CI: 0.22−1.65; men aRRR 0.64, 95% CI: 0.27−1.52) than those with an SA due to cancer (Table 2 and 3). In men, this LMPP was the third most prevalent (11.4%), with almost 100% of men being retired by the end of follow-up. Women showed a similar LMPP, but in contrast, it was the least frequent pattern (Figure 2). Both men and women in these patterns were over 55 years and worked in a private company (Table 1).

## DISCUSSION

This study confirms our hypothesis that workers with an SA due to cancer were more likely to retire, receive PD benefits or die than those without a previous SA. These findings are consistent after adjustment by several employment and working conditions.

Our results on retirement in workers with an SA due to cancer agree with those of previous literature. Workers very close to retirement age who are diagnosed with cancer may choose to retire [20]. Cancer treatments are still, in general, highly aggressive and leave some survivors with long-term health problems that may make them unable to carry out their prior employment, requiring them to live on PD. For example, a systematic review found that long-term survivors were less likely to be working than people without cancer [21]. Some workers experience less severe side effects from treatments, but require longer SA spells for recovery and readaptation to be able to work. In this regard, the social security system plays a major role by setting a maximum amount of time on SA that does not always suit cancer survivors’ needs. These results question the effectiveness of the Spanish social security system in maintaining workers in the labour market during their working life and, when they are ill, to guarantee their income through benefits. In this case, after cancer treatment, some workers could continue working, but require more flexible SA schemes to recover from the range of effects produced by the cancer. Another explanation could be that workers who have had a serious health problem may find it more difficult to maintain the pace of a normal full-time job in the long term. However, they might also encounter discrimination – including hiring discrimination, harassment, job reassignment, job loss, and limited career advancement– due to their health problem [22]. These results are also coherent with the well-known healthy worker effect [23].

When we compared the probability of having a stable future working life to other patterns, we found unexpected results in workers with a SA due to cancer. In men, in general, they were less likely to be in unstable and early retirement, or in PD trajectories than those with SA due to other causes. We hypothesised that future instability could lie in the fact that SA due to other causes were highly represented by diseases of the musculoskeletal system and connective tissue, and these types of diagnoses are more prevalent among manual workers [24,25], who normally have more precarious jobs characterised by temporality and insecurity in Catalonia [26] and in the whole of Spain [27]. Few studies have assessed RTW and long-term working trajectories regarding PD or early retirement after all-cause SA comparing them with those with an SA due to cancer. A study carried out in Norway with a 10-year follow-up that examined long-term SA due to all diagnoses found that 32% of workers with SA due to any diagnoses had low attachment labour market trajectories, consisting of part-time work, recurrence of SA, unemployment or PD [28]. In women, we observed similar tendencies of unstable future working life among those with an SA due to other causes.

This study identified differences in employment patterns by sex. Only in women, we found an LMPP of temporary employment. This LMPP was more probable among workers with an SA due to cancer than among workers with no SA. In Spain, temporary and part-time employment is more prevalent among women due to huge gender inequities in the Spanish labour market [29]. A similar study carried out in northern countries using the same methodology, but examining all-cause SA, found no differences by sex [30], whereas another study found that women were more prone to be in less stable trajectories with part-time work and SA recurrence than men [28]. It is likely that this gender-based temporariness is exacerbated by cancer.

We also found differences by sex in retirement. In men, the difference in the likelihood of retirement among workers with an SA due to cancer compared with those without an SA was larger than that in women. We found a percentage of male workers that started to retire early, and this percentage increased steadily during the follow-up period. Also, retirement pattern was twice as prevalent as that found in women. This might be due to differences in ease of access to retirement for several reasons. Firstly, as in all countries, a minimum period of pension contributions is a prior condition for retirement or early retirement. Due to reproductive work and motherhood, women have often contributed fewer years to the social security system, and for early retirement, workers must have contributed for at least 35 years [31,32]. Secondly, men are generally in better paid jobs, and since the amount of the early retirement pension depends on the workers’ previous regulatory basis, men have better early retirement pensions, making retirement more appealing [29]. In addition, some companies have early retirement policies if the worker voluntarily wants to retire and does not meet social security requirements, and these policies are more accessible to men than to women [33], probably because they exist in male dominated sectors. Lastly, in our study, the sample of men was older than that of women and, throughout Spain, to retire before the retirement age, the worker must be a maximum of 2 years below this age to get full pension benefits.

The main limitation of the present study is the lack of information on cancer other than the SA diagnosis for the period 2012-2015. Hence, we were unable to take into consideration the effect of clinical features (i.e., type of treatment, stage of cancer, effects of health status before 2012). In addition, we managed time-varying covariates by assigning workers to the category in which they spent most of their time during the follow-up. Also, self-employed were not included in the study, because they did not have access to SA benefits during the study period. Furthermore, the methodology applied to employment trajectories involved an algorithmic data-driven approach that classified individuals according to similar characteristics. Consequently, some resulting groups had a small number of observations which should be interpreted cautiously.

The primary strength of this study, in terms of occupational health relevance, as well as from a clinical perspective, is the use of administrative data with information on social security benefits and an extensive time window of follow-up with information on labour market states, which is needed to obtain a sufficient overview of the RTW process. The size of the database allowed us to match our workers with SA due to cancer to two comparison groups by age, sex, and follow-up time. This matching allowed us to compare the working life of cancer survivors with that of the general working population with or without SAs, enabling us to account not only for the disease, but also for the effect of SA. The diagnoses causing SAs were medically certified by primary care doctors rather than self-reported, enhancing the validity of our results [34].

## CONCLUSIONS

Most workers with an SA due to cancer have a future working trajectory in employment. However, probably due to the disease and the adverse effects of treatment, they are more likely to retire, receive a PD benefit or die than their counterparts of the same age and sex. This study represents a step towards a deeper understanding of the consequences of cancer on working life as it captures all possible labour outcomes and their chronological onset on workers’ life. However, more studies are needed to address questions that remain unanswered in the light of our results such as the whether there any differences between cancer diagnoses, stages, or treatments regarding future working trajectories.

Nevertheless, for the time being, action should be taken to regulate programmes in the workplaces that consider the needs of workers who have survived cancer, many of which are common to all diagnoses, so that these workers may remain working when possible and desired.

## Data Availability

All data produced in the present study are available upon reasonable request to the authors

## ACKNOWLEDGEMENTS

This work would not have been possible without the Spanish National Social Security Institute and the Catalonian Institute for Medical Evaluations.

## AUTHOR CONTRIBUTIONS

All listed authors fulfil authorship criteria. A.A.G., L.S. and F.G.B. participated in the study conception and design. A.A.G. and L.S. performed the main data management and analysis and interpreted the data. A.A.G. drafted the first version of the manuscript with close help from L.S. F.G.B. and L.S. made subsequent revisions to the manuscript. All authors revised the final version, agreed with the text and findings, and approved this final version. The corresponding author certifies that all listed authors meet authorship criteria. A.A.G. is the guarantor.

## FUNDING

The study was financed by the State Plan for Investigation, Development, and Innovation, 2021–2023, by the Health Institute Carlos III-FIS_FEDER—Subdirection General of Evaluation and Promotion of Investigation (Grants FIS PI20/00101 and FIS PI17/00220). The study was also supported by the Spanish Society of Epidemiology and the CIBER of Epidemiology and Public Health.

## COMPETING INTERESTS

The authors declare no competing interests.

## Notes

### Competing Interest Statement

The authors have declared no competing interest.

### Funding Statement

This study was funded by Health Institute Carlos III- FIS_FEDER

### Author Declarations

This study was approved by the Parc de Salut Mar Ethics Committee in Barcelona (Research Protocol no. 2020/9119) and was exempted from informed consent requirements owing to its registry-based design

